# Cognitive ability in offspring conscripts and cardiovascular disease risk in extended family members: assessing the impact of modifiable risk factors on familial risk

**DOI:** 10.1101/2024.06.03.24308018

**Authors:** María Fernanda Vinueza-Veloz, Paul Remy Jones, Marte Karoline Råberg Kjøllesdal, Huong Thu, David Carslake, Øyvind Næss

## Abstract

**Background and aim:** Previous studies have demonstrated an inverse association between cognitive ability (CA) and risk of cardiovascular diseases (CVD). This study aims to investigate the associations between CA in offspring and CVD mortality in relatives of the parental generation (i.e., parents, aunts/uncles (A/U), and A/U partners), and assess the role of modifiable risk factors on these associations.

**Methods:** This longitudinal study included nearly 3 million adults who were followed up from age 45 until death. Data for participants were obtained through the linkage of various Norwegian surveys and registries. Hazard ratios (HR) for CVD mortality among the parental generation in relation to offspring CA were estimated using Cox proportional hazard regression.

**Results:** One standard deviation (SD) unit increase in CA was associated with a reduction in CVD mortality among mothers (HR: 0.77, 95%CI [0.74, 0.81]); fathers (0.83, [0.81, 0.86]); A/U (0.91, [0.87, 0.94]); and A/U partners (0.91, [0.89, 0.94]). Adjusting the models for CVD risk factors in the parental generation attenuated all associations (mothers-HR: 0.91, 95%CI [0.87, 0.96]; fathers: 0.93, [0.91, 0.96]; A/U: 0.99, [0.96, 1.03]; A/U partners: 0.98, [0.95, 1.01]).

**Conclusions:** We observed an inverse association between offspring CA and CVD and all-cause mortality in various familial relationships. Our findings suggest the existence of factors shared among relatives that explain familial risk to suffer lower CA and higher CVD mortality. A significant portion of the association between CA and CVD mortality in all familial relationships was explained by modifiable risk factors in relatives of the older generation.

**Highlights:**

- A lower cognitive ability might drive inequalities in cardiovascular disease (CVD).
- Shared environments explain most of the association between cognitive ability and CVD.
- The role of genetic factors in this relationship has probably been overestimated.

## Introduction

Cognitive ability (CA) measured in childhood and late-adolescence has been associated with all-cause mortality risk, including death due cardiovascular diseases (CVD), as well as many other health outcomes in adulthood (1,2). Several general non-exclusive explanations have been suggested for the association between CA and physical health and mortality (3). First, childhood CA may be picking up bodily insults including poor maternal health, intrauterine growth retardation, malnutrition, and adverse socio-economic circumstances. Secondly, it may be a predictor of healthy behaviour and capacity to enter safe environments. Thirdly, CA may be a marker of general bodily system integrity. Evidence for the latter comes from the fact that CA is associated with a range of different causes of death, each with their own set of risk factors (4,5). Similarly, genetically informed studies have showed genetic correlation between CA and various health outcomes acting through common genetic (pleiotropic) mechanisms (6,7).

For CVD, the role of modifiable risk factors is crucial. Additionally, CA has increasingly been recognized as significant in contributing to health inequalities in populations with high income and education levels (8). For both CA and social inequalities, it has important public health and policy interest to know if targeting risk factors will be sufficient to improve population level incidence of CVD and related non-communicable diseases with the same risk factors as CVD (9). The association between childhood and adolescent CA and later life CVD may origin from latent biological traits that act independently from established risk factors for CVD. For example, CA has been related to metabolic syndrome, major depression, and cumulative allostatic load (10–12).

Yet, at population level 90% of attributable risk in myocardial infarction between countries can be accounted for by a few key established CVD risk factors including smoking, unfavourable levels of blood lipids, and high blood pressure (13). Most of the social inequalities in CVD is also attributable to these risk factors, especially when they are measured repeatedly over the life course or when absolute measures of risk are used (14,15). However, to what extent the association between early life CA and CVD is explained and modifiable by risk factors is not well known yet. The factors that determine the association between early life CA and later CVD risk, are probably closely linked with family factors (3). One approach to investigate important components of family factors on the association between CA and later life CVD is to relate CA in offspring with CVD in the parental generation and investigate the role of modifiable risk factors (16).

Previously, the role of family factors has been studied applying parent–offspring designs by comparing CVD mortality in mothers and fathers in relation to exposures such as birth weight or body mass index in their offspring (17,18). In such designs measuring exposures in the offspring is used as proxy of measuring exposures in the parents. Comparing maternal vs. paternal associations helps to understand mother-specific contributions including, intrauterine factors as well as mother’s education, nutrition and lifestyle (19). In addition to the parent–offspring design, it may be valuable to investigate CVD mortality in extended family members with known genetic relatedness to the offspring and who are less likely to share environment with them, such as aunts/uncles (A/U) and A/U partners (18).

The objective of the present study was first to investigate the associations between offspring CA and risk of CVD in the parental generation with different degrees of genetic relatedness (parents, A/U, A/U partners), and secondly to assess the role of established modifiable risk factors.

## Methods

### Data sources and study population

Data were obtained by linking health surveys with various registries: The Age 40 Program, The Counties Study, The Cohort Norway (CONOR), Norwegian Armed Forces Heath Registry, Norwegian Population Registry, and generational data from the Norwegian Family Based Life Course Study (NFLC). Additional sources included the National Educational Registry, and the Cause of Death Registry (20–23). The study population comprised Norwegian male conscripts, born between 1948 and 1994 who were included in the Armed Forces’ personnel registry, along with their family members.

Conscripts were linked to their mothers (n = 614 650) and fathers (n = 561 889), who were then linked to their siblings, i.e. the conscript’s A/U (n = 1 016 127). A/Us were further linked to their first spouses (A/U partners) as recorded in the Norwegian Population Registry (n = 869 711). Detailed information on how the sample was delimited can be seen in Figure 1. A sub-sample of the full sample included all relatives with measured CVD risk factors: n mothers, 235 475; n fathers, 202 805; n A/U, 359 143; n A/U partners, 325 478 (Figure 1).

**Figure 1.**
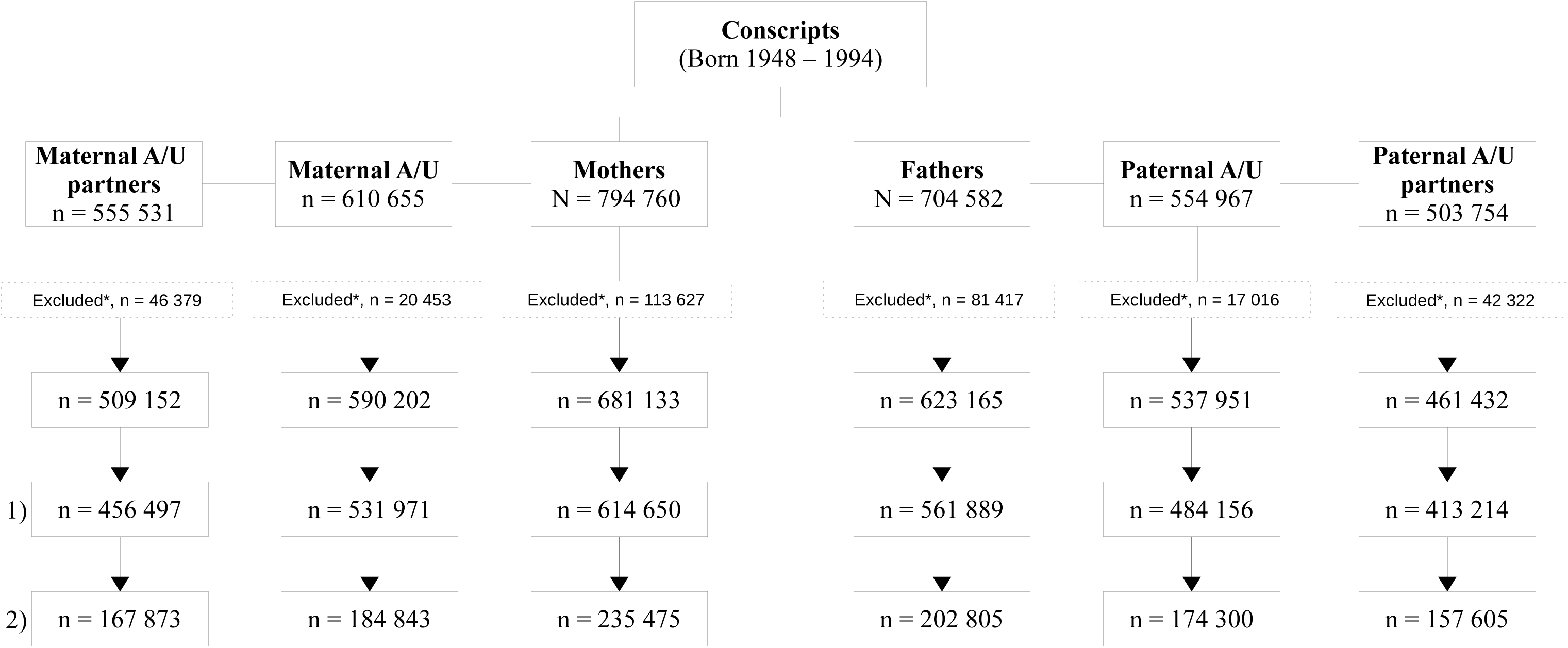
Chart-flow of sample selection. Norwegian conscripts born between 1948 and 1994 were linked to their mothers and fathers, who were linked to their corresponding siblings and them to their partners. *Abbreviations and symbology: A/U, aunts/uncles; n or N, number; *, Exclusion criteria: relatives of conscripts who were dead before the age of 45 or born before 1939; 1), full-sample with full information on sex, birth year and level of education; 2) sub-sample, with full information on sex, birth year, level of education and cardiovascular risk factors (see Methods)*.

### Outcome

Data on CVD and all-cause mortality were retrieved from the Norwegian Cause of Death Registry. CVD mortality was defined using codes 390-459 from the International Classification of Diseases ninth revision (ICD-9) and I00-I99 from the tenth revision (ICD-10), corresponding to diseases of the circulatory system.

### Exposure

Data corresponding to CA of conscripts were retrieved from The Norwegian Armed Forces Heath Registry. CA was assessed through three different, time-limited sub-tests: Arithmetic (25 minutes), Word Similarities (8 minutes) and Figures (20 minutes) (20). The Arithmetic and Word Similarities tests were similar to those found in the Wechsler Adult Intelligence Scale (WAIS) (24). The Figures test was designed to resemble Raven’s Progressive Matrices (25). Scores from each of the three tests were standardized into normally distributed F-scores (mean = 50, standard deviation = 20) and summed to yield a combined measure of general CA, reported as a stanine score from 1 to 9 (20). A stanine score of five corresponds to an IQ of 100 in WAIS, with each stanine unit deviation from five representing an increment or decrement of 7.5 IQ units. CA was analysed as continuous in Cox models, but categorized into low (stanine score 1 – 3), medium (4–6), and high (7–9) for descriptive purposes.

### Covariates

Education level and CVD risk factors were measured in relatives of the parental generation. Data on educational level (i.e., years of education) was obtained from the National Educational Registry. Educational level was analysed as a continuous covariate in Cox models, but categorized into low (≤ 9 years) and high (>9 years) for descriptive purposes. Analysed CVD risk factors included body mass index in kg/m2 (BMI), non-fasting serum total cholesterol (mmol/L), blood pressure (mmHg), and self-reported smoking status (yes/no). Details on measurement techniques can be consulted in previous publications (21–23).

### Statistical analysis

To estimate the hazard ratios (HR) of CVD mortality in the parental generation by the level of offspring CA, we implemented Cox proportional hazard regression models with age as underlying time variable. Offspring CA stanine scores were converted to standard deviation (SD) units (1 SD = 1.75 stanine units = 13.13 IQ units) and analysed as continuous. Separate models were implemented for mothers, fathers, combined maternal and paternal A/U, and combined maternal and paternal A/U partners. Parents, A/Us, and A/U partners were followed up from age 45 until the earliest of (i) death from any cause, (ii) emigration, or (iii) administrative censoring (31 December 2020). The proportional hazard assumption was checked by visual inspection of residuals, and no violations were identified. Since there were several conscript observations per parent, we estimated cluster-robust standard errors within family membership. Models were adjusted for birth year as a continuous variable in the parental generation, in addition to sex for A/U and A/U partners (model 1), covariates of model 1 plus level of education (model 2), covariates of model 1 plus CVD risk factors (model 3), and covariates of model 1 plus education and CVD risk factors (model 4). CA was also analysed as categorical variable, where stanine 1 was compared to stanines 2 to 9. We also introduced an interaction term between offspring CA and parental sex to check differential effects for mothers and fathers.

## Results

The mean CA of conscripts linked to mothers was 5.09 (SD, 1.76) with a median of 5, and their median birth year was 1979 (range: 1948 to 1994). All relatives were born between 1940 and 1975. At the end of the follow-up period, family members linked to offspring with high CA were older and more educated than those linked to offspring with medium or low CA (Table 1). Fathers were slightly older and more educated than mothers (Table 1). A dose-dependent relationship was observed, whereby family members of offspring with high CA had a lower percentage of CVD and all-cause deaths than those of offspring with medium or low CA. Furthermore, there was a lower percentage of deaths among mothers compared to fathers, A/U, and A/U partners (Table 1). Mothers had lower BMI, lower levels of non-fasting cholesterol, systolic blood pressure (SBP), and smoked less than fathers (Table 2).

**Table 1.**
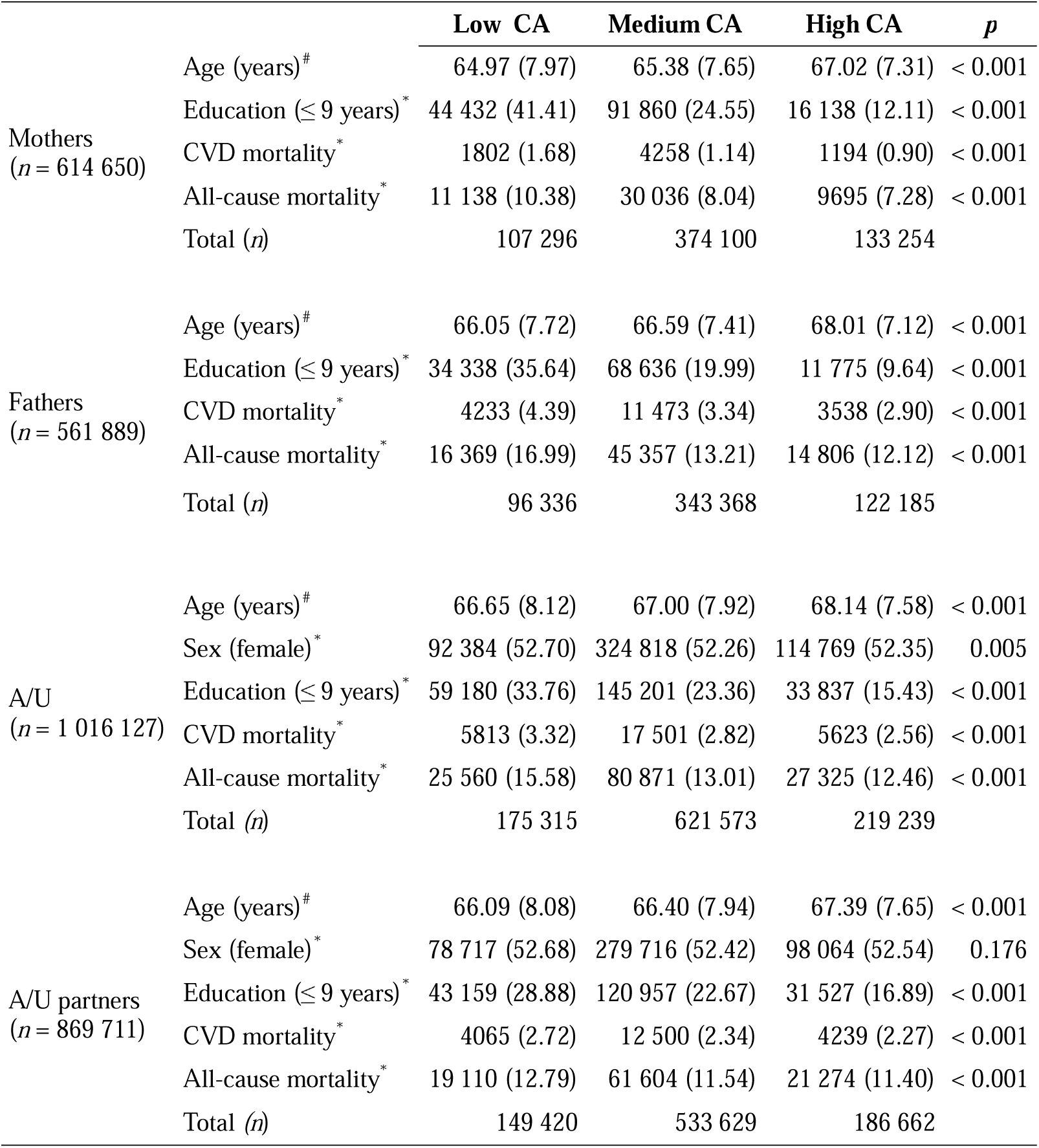
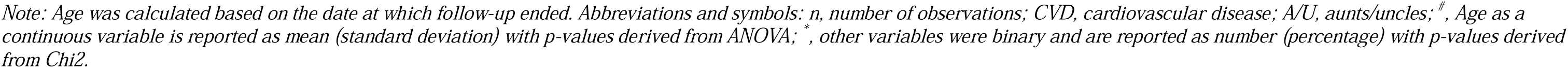
Characteristics of the full-sample according to offspring cognitive ability. Stanine scores of cognitive ability (CA) were categorized into three groups: low (1–3), medium (4–6) and high (7–9).

**Table 2.** Description of cardiovascular (CVD) risk factors in the sub-sample of relatives with measured CVD risk factors. Stanine scores of cognitive ability (CA) were categorized into three groups: low (1–3), medium (4–6) and high (7–9).

|  |  | Low CA | Medium CA | High CA | *p* |
| --- | --- | --- | --- | --- | --- |
| Mothers<br>(*n* = 235 475) | BMI (kg/m<sup>2</sup>)<sup>#</sup> | 24.37 (3.85) | 24.45 (3.84) | 24.09 (3.61) | < 0.001 |
|  | Non-fasting cholesterol (mmol/L)<sup>#</sup> | 5.57 (1.15) | 5.46 (1.00) | 5.37 (0.97) | < 0.001 |
|  | SBP (mmHg)<sup>#</sup> | 127.07 (14.38) | 125.85 (14.05) | 125.08 (13.71) | < 0.001 |
|  | Smoking daily (yes)<sup>*</sup> | 18 067 (49.91) | 54 260 (39.07) | 15 550 (28.78) | < 0.001 |
|  | Total (*n*) | 37 565 | 142 603 | 55 307 |  |
| Fathers<br>(*n* = 202 805) | BMI (kg/m<sup>2</sup>)<sup>#</sup> | 26.00 (3.33) | 25.78 (3.13) | 25.47 (3.00) | < 0.001 |
|  | Non-fasting cholesterol (mmol/L)<sup>#</sup> | 5.93 (1.13) | 5.84 (1.10) | 5.77 (1.08) | < 0.001 |
|  | SBP (mmHg)<sup>#</sup> | 81.74 (10.12) | 81.21 (9.98) | 81.00 (9.87) | < 0.001 |
|  | Smoking daily (yes)<sup>*</sup> | 14 549 (47.23) | 46 870 (38.52) | 13 707 (29.90) | < 0.001 |
|  | Total (*n*) | 31 720 | 124 395 | 46 690 |  |
| A/U<br>(*n* = 359 143) | BMI (kg/m<sup>2</sup>)<sup>#</sup> | 25.30 (3.83) | 25.13 (3.68) | 24.90 (3.61) | < 0.001 |
|  | Non-fasting cholesterol (mmol/L)<sup>#</sup> | 5.73 (1.10) | 5.68 (1.12) | 5.64 (1.06) | < 0.001 |
|  | SBP (mmHg)<sup>#</sup> | 79.46 (10.58) | 79.20 (10.51) | 79.05 (10.44) | < 0.001 |
|  | Smoking daily (yes)<sup>*</sup> | 12 230 (43.69) | 38 960 (38.19) | 12 150 (33.11) | < 0.001 |
|  | Total (*n*) | 60 262 | 219 317 | 79 564 |  |
| A/U partners<br>(*n* = 325 478) | BMI (kg/m<sup>2</sup>)<sup>#</sup> | 25.20 (3.78) | 24.97 (3.63) | 24.79 (3.54) | < 0.001 |
|  | Non-fasting cholesterol (mmol/L)<sup>#</sup> | 5.69 (1.09) | 5.65 (1.15) | 5.61 (1.07) | < 0.001 |
|  | SBP (mmHg)<sup>#</sup> | 79.03 (10.37) | 78.73 (10.43) | 78.60 (10.36) | < 0.001 |
|  | Smoking daily (yes)<sup>*</sup> | 11 673 (44.66) | 37 279 (39.59) | 11 697 (35.09) | < 0.001 |
|  | Total (*n*) | 55 154 | 199 014 | 71 310 |  |

Abbreviations and symbols: *n*, number of observations; BMI, body mass index; SBP, systolic blood pressure; A/U, aunts/uncles; <sup>#</sup>, continuous variables are reported as mean (standard deviation) with *p*-values derived from ANOVA; <sup>\*</sup>, smoking was binary and reported as number (percentage) with *p*-values derived from Chi2.

### Cardiovascular disease mortality

Among mothers, fathers, A/U and A/U partners, CVD mortality was inversely associated with offspring CA. Hazard ratios (HRs) for CVD mortality were slightly closer to the null among fathers than among mothers. While mothers had a crude 23% reduction in CVD mortality per standard deviation (SD) unit increase in CA (1.75 stanine units), the equivalent figure in fathers was 17% (Table 3, Figure 2). The CA-CVD association was stronger for mothers than for fathers (p = 0.009). The reduction in hazard for CVD mortality per SD unit increase in CA was also present, although attenuated, in A/U (9%) as well as A/U partners (9%) (Table 3, Figure 2).

**Figure 2.**
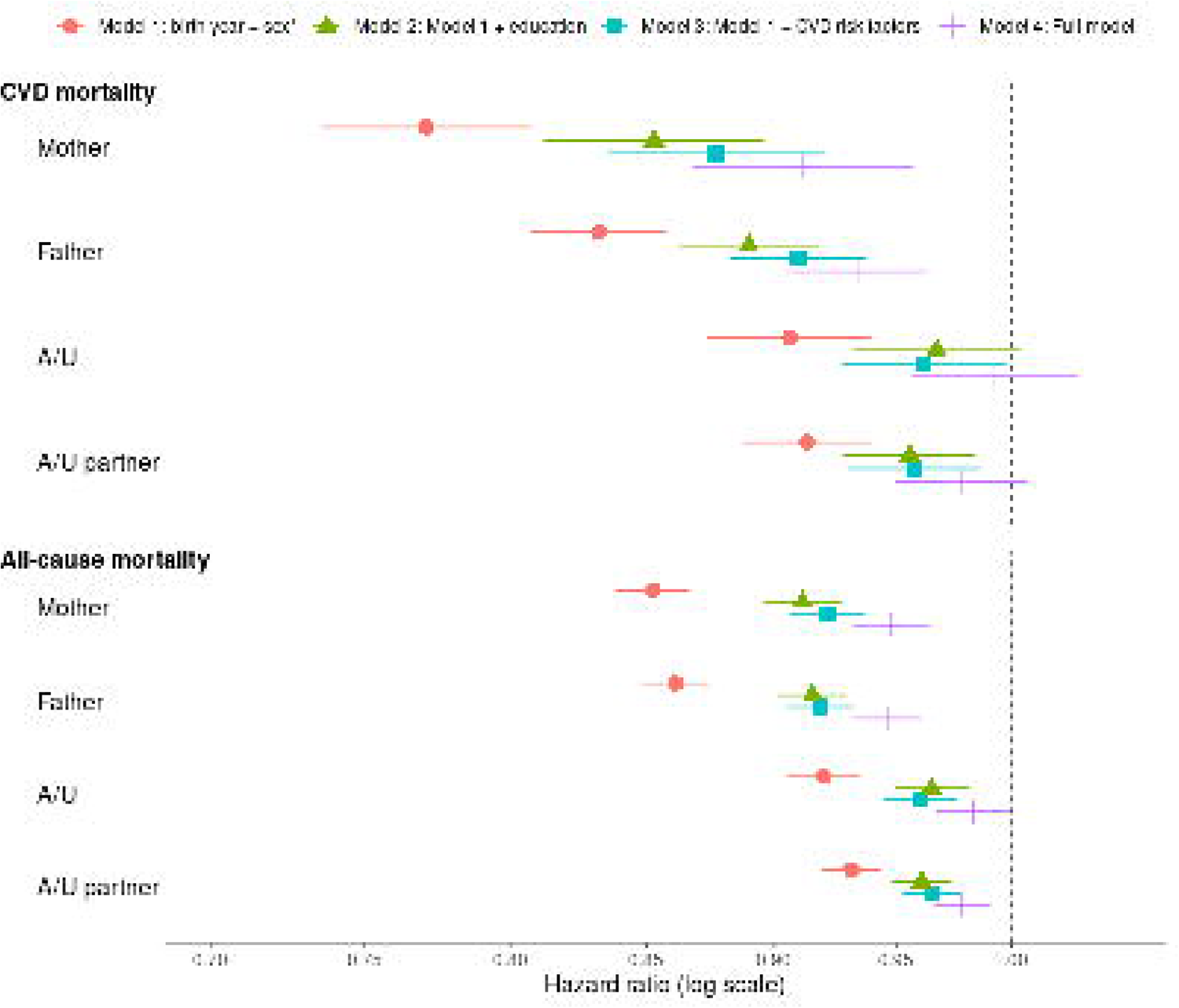
Hazard ratios for cardiovascular and all-cause mortality among the parental generation per standard deviation (SD) unit of offspring cognitive ability in the sub-sample of relatives with measured CVD risk factors. Each SD unit is equal to 1.75 stanine or 13.13 IQ units. Hazard ratios (HR) and their corresponding 95% confidence intervals are shown for mothers, fathers, aunts/uncles (A/U) and A/U partners. In model 1, HR were adjusted for birth year and sex of the parental generation (A/U and A/U partners). In model 2, HR were adjusted for covariates of model 1 plus level of education in the parental generation. In model 3, HR were adjusted for covariates of model 2 plus CVD risk factors in the parental generation.

**Table 3.**
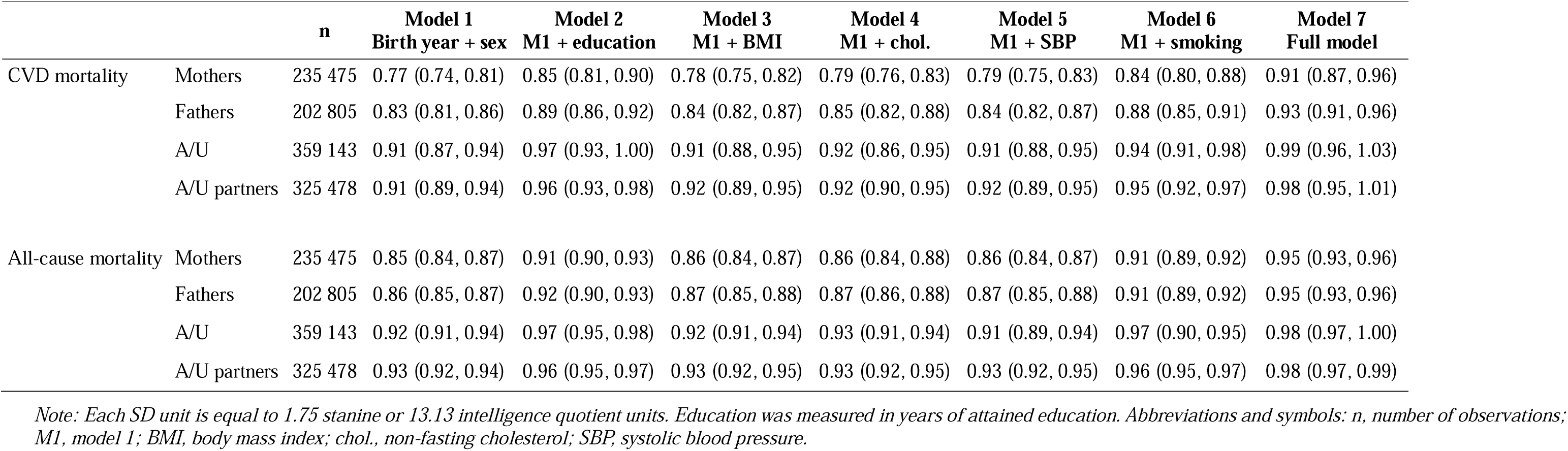
Hazard ratios for cardiovascular (CVD) and all-cause mortality among the parental generation per standard deviation (SD) unit of offspring cognitive ability (sub-sample). Hazard ratios (HR) and their corresponding 95% confidence intervals are shown for mothers, fathers, aunts/uncles (A/U) and A/U partners.

|  |  | n | Model 1<br>Birth year + sex | Model 2<br>M1 + education | Model 3<br>M1 + BMI | Model 4<br>M1 + chol. | Model 5<br>M1 + SBP | Model 6<br>M1 + smoking | Model 7<br>Full model |
| --- | --- | --- | --- | --- | --- | --- | --- | --- | --- |
| CVD mortality | Mothers | 235 475 | 0.77 (0.74, 0.81) | 0.85 (0.81, 0.90) | 0.78 (0.75, 0.82) | 0.79 (0.76, 0.83) | 0.79 (0.75, 0.83) | 0.84 (0.80, 0.88) | 0.91 (0.87, 0.96) |
|  | Fathers | 202 805 | 0.83 (0.81, 0.86) | 0.89 (0.86, 0.92) | 0.84 (0.82, 0.87) | 0.85 (0.82, 0.88) | 0.84 (0.82, 0.87) | 0.88 (0.85, 0.91) | 0.93 (0.91, 0.96) |
|  | A/U | 359 143 | 0.91 (0.87, 0.94) | 0.97 (0.93, 1.00) | 0.91 (0.88, 0.95) | 0.92 (0.86, 0.95) | 0.91 (0.88, 0.95) | 0.94 (0.91, 0.98) | 0.99 (0.96, 1.03) |
|  | A/U partners | 325 478 | 0.91 (0.89, 0.94) | 0.96 (0.93, 0.98) | 0.92 (0.89, 0.95) | 0.92 (0.90, 0.95) | 0.92 (0.89, 0.95) | 0.95 (0.92, 0.97) | 0.98 (0.95, 1.01) |
| All-cause mortality | Mothers | 235 475 | 0.85 (0.84, 0.87) | 0.91 (0.90, 0.93) | 0.86 (0.84, 0.87) | 0.86 (0.84, 0.88) | 0.86 (0.84, 0.87) | 0.91 (0.89, 0.92) | 0.95 (0.93, 0.96) |
|  | Fathers | 202 805 | 0.86 (0.85, 0.87) | 0.92 (0.90, 0.93) | 0.87 (0.85, 0.88) | 0.87 (0.86, 0.88) | 0.87 (0.85, 0.88) | 0.91 (0.89, 0.92) | 0.95 (0.93, 0.96) |
|  | A/U | 359 143 | 0.92 (0.91, 0.94) | 0.97 (0.95, 0.98) | 0.92 (0.91, 0.94) | 0.93 (0.91, 0.94) | 0.91 (0.89, 0.94) | 0.97 (0.90, 0.95) | 0.98 (0.97, 1.00) |
|  | A/U partners | 325 478 | 0.93 (0.92, 0.94) | 0.96 (0.95, 0.97) | 0.93 (0.92, 0.95) | 0.93 (0.92, 0.95) | 0.93 (0.92, 0.95) | 0.96 (0.95, 0.97) | 0.98 (0.97, 0.99) |

*Note: Each SD unit is equal to 1.75 stanine or 13.13 intelligence quotient units. Education was measured in years of attained education. Abbreviations and symbols: n, number of observations; M1, model 1; BMI, body mass index; chol., non-fasting cholesterol; SBP, systolic blood pressure.*

A substantial proportion of the hazard for CVD mortality was accounted for by level of education in the parental generation, as all HRs attenuated after adjustment for it (Table 3, Figure 2). Among mothers, the reduction in hazard for CVD mortality per SD unit increase in CA decreased from 23% to 15%, for fathers from 17% to 11%, for A/U from 9% to 3%, and for A/U partners from 9% to 4%. Both the full- and the sub-sample displayed very similar patterns of associations (Supplementary table 1).

Adjustment for CVD risk factors in the parental generation attenuated the effect size even more than the adjustment for education, rendering it close to the null among A/U and A/U partners (Table 3, Figure 2). In the fully adjusted model, the CA-CVD association exhibited a similar strength for mothers and fathers (p = 0.095). Among mothers, the reduction in hazard for CVD mortality per SD unit increase in CA decreased from 23% to 12%, for fathers from 17% to 10%, for A/U from 9% to 1%, and for A/U partners from 9% to 2% (Table 3, Figure 2).

### All-cause mortality

Among mothers, fathers, A/U, and A/U partners, all-cause mortality was inversely associated with offspring CA. Generally, the reduction in hazard for all-cause mortality per SD unit increase in CA was similar between mothers and fathers, and between A/U and A/U partners (Table 3). Mothers and fathers experienced a crude 14 to 15% lower all-cause mortality per SD unit increase in CA. The CA-all-cause mortality association exhibited a similar strength for mothers and fathers (p = 0.405). The reduction in hazard for all-cause mortality per SD unit increase in CA was also present, albeit attenuated, in A/U (8%) and A/U partners (7%) (Table 3).

A substantial part of the hazard of all-cause mortality was accounted for by level of education of the parental generation (Table 3). Among mothers, the reduction in hazard for all-cause mortality per SD unit increase in CA decreased from 15% to 9%, for fathers from 14% to 8%, for A/U from 8% to 3%, and for A/U partners from 7% to 4%. Both the full- and the sub-sample demonstrated similar pattern of associations between offspring CA and all-cause mortality (Supplementary table 1).

Adjustment for CVD risk factors in the parental generation slightly attenuated the effect size and produced similar estimates to those for education (Table 3, Figure 2). In the fully adjusted models, the CA-all-cause mortality association exhibited a similar strength for mothers and fathers (p = 0.095).

Analyses of CA as categorical (see Methods section) suggested a dose-response association with CVD and all-cause mortality, with HRs further from the null for the highest stanine values (2 to 9) relative to stanine 1. This was the case in the full sample and in the sub-sample with measured CVD risk factors (Supplementary table 2, 3 and 4).

## Discussion

The aim of this study was to investigate associations between offspring CA and CVD in the parental generation, and to assess the impact that modifiable CVD risk factors have. We found that higher offspring CA was associated with lower CVD mortality among mothers, fathers, A/U, and non-genetically related family members (i.e., A/U partners). Both level of education and CVD risk factors accounted for a significant portion of the CA-CVD association across all studied familial relationships (offspring-parent, offspring-A/U and offspring-A/U partners).

### Associations within individuals and between parents and offspring

The results of our study confirm the inverse association between CA and mortality previously described in a Swedish study with an analogous design, and in others analysing within-person associations (2,26–29). Similarly to the Swedish study, we observed that offspring-mother estimates were stronger than offspring-father estimates in the unadjusted models (27). However, after accounting for level of education and/or CVD risk factors the difference between maternal and paternal estimates diminished. The difference between maternal and paternal estimates in the unadjusted models may be explained by mother specific factors acting through health behaviours such as smoking and nutrition, that affect offspring pre- or postnatally (30,31).

### Associations between offspring and aunts/uncles

The association between offspring CA and CVD mortality in A/Us points toward mechanisms beyond those shared within the nuclear family. Sometimes, alleles from the genotype of the parent are not passed on to the offspring yet still influence the phenotype of the offspring through indirect genetic effects. This mechanism has been shown to be influential for socioeconomic position and education, and is also likely to be so for CA (32). To what extent this mechanism plays a role across traits, i.e. between CA or education and risk of CVD is yet still not well known. Such effects may stretch beyond first degree relatives as dynastic effects. Evidence on the role of these come from family GWAS and extended family studies (33). The attenuation in a large within sibling GWAS consortium was particularly pronounced for behavioural traits. Even though CA was not included in these family studies, it is likely that CA may follow similar pattern as education and health behaviour.

### Associations with non-genetically related partners of aunts and uncles

The associations with A/U partners are consistent with findings from various studies showing pronounced levels of assortative mating within traits like CA and education. Assortative mating is greater for CA than for other behavioural or physical traits (34). Assortative mating by CA will give children with higher CA than average. This increases additive genetic variance of CA resulting in accumulation across generations. Recent evidence in distant family members using polygenic indices of several indicators including education showed greater genetic similarity between distant relatives than close ones, which is consistent with the comparatively large association with A/U partners (35). Assortative mating across traits also introduces an alternative explanation to pleiotropy. In UK biobank widespread cross-trait assortative mating, including education and CA with a range of physical traits, indicates that genetic correlations might be inflated (36).

### Implications

Our findings suggest that the familial components of the association between CA in early life and CVD is to a large extent explained by modifiable risk factors. This is important for public health prevention of CVD because it underlines the importance of these risk factors and their associated health behaviour. The aim here was not to disentangle the role of different mechanisms. It is a matter of ongoing and future research to sort out the relative influence of these for familial associations. This is limited by paucity of family genetic data. The sibling GWAS demonstrated pronounced familial effects providing empirical evidence for several theoretical mechanisms yet could not point to which had the strongest impact (33). The present state of knowledge in this rapidly evolving field and evidence from our study suggest that CA and modifiable risk factors are deeply embedded in the family of origin and that several mechanisms may play a role.

Familial associations probably originate through an interplay between genetic and environmental factors involving several of the previously presented mechanisms. Even if these associations may be strongly related to family factors and present as liability in genetically informed studies, the levels people end up havering of these risk factors are not determined by these family factors as they are highly modifiable after birth. This has been an important premise for the prevention strategy for children with familial hypercholesterolemia (37). As CA is strongly linked to social inequalities, our results suggest that prevention of inequalities in CVD may benefit from proper attention to health behaviour in children.

### Strengths and limitations

This is one of few population-based studies that includes data from nationwide registers and health surveys spanning two generations. Moreover, we have described the association between CA and CVD mortality among relatives with various levels of genetic relatedness. Additionally, our sample is well-powered, providing reasonably precise estimates. However, there are some limitations to consider. Notably, very few women underwent CA testing, resulting in the exclusion of female offspring and their relatives from our analysis. Furthermore, CVD risk factors were not measured across the entire sample, but only among individuals who voluntarily participated in national and regional health surveys, potentially introducing selection bias. Nevertheless, as the estimates in the full and the sub-sample for the associations between CA and CVD were comparable in size, the influence of selection bias is expected to be limited. The associations in parents were not fully explained by risk factors, which means we cannot rule out other mechanisms.

## Conclusions

A significant portion of associations between CA in offspring and CVD in separate family relationships were explained by modifiable risk factors. This suggests the familial part of the CA CVD association is primarily driven by risk factors.

## Ethics approval

The study protocol was approved by the Regional Committees for Medical Research Ethics South East Norway (REK 2012/827).

## Data availability

Stata scripts used for the analyses in this article are available upon request. Individual-level data can be requested to Statistics Norway’s registry data.

## Author contributions

The first author of this work declares that all authors have contributed and work on the development of it as follows. MFVV and ØN: Conception and design, analysis, and interpretation of data, drafting of the article, critical revision for important intellectual content, final approval of the version to be published, agreement to be accountable for all aspects of the work. PRJ, MKRK, HT, DC: Interpretation of data, critical review for important intellectual content, final approval of the version to be published, agreement to be accountable for all aspects of the work.

## Funding

ØN, MFVV, PRJ and HNT were funded by The Research Council of Norway, Project Inequalities in non-communicable diseases: Indirect selection or social causation (Project number 287347). DC works in a unit funded by the UK Medical Research Council (MC_UU_00011/1) and by the University of Bristol.

## Conflict of interest

None to declare.

## Notes

### Competing Interest Statement

The authors have declared no competing interest.

### Funding Statement

ON, MFVV, PRJ and HNT were funded by The Research Council of Norway, Project Inequalities in non-communicable diseases: Indirect selection or social causation (Project number 287347). DC works in a unit funded by the UK Medical Research Council (MC_UU_00011/1) and by the University of Bristol.

## References

1. Dobson KG, Chow CHT, Morrison KM, Van Lieshout RJ. Associations Between Childhood Cognition and Cardiovascular Events in Adulthood: A Systematic Review and Meta-analysis. Canadian Journal of Cardiology. 2017 Feb 1;33(2):232–42.

2. Hemmingsson T, Essen J v., Melin B, Allebeck P, Lundberg I. The association between cognitive ability measured at ages 18-20 and coronary heart disease in middle age among men: A prospective study using the Swedish 1969 conscription cohort. Social Science and Medicine. 2007 Oct;65(7):1410–9.

3. Deary IJ, Hill WD, Gale CR. Intelligence, health and death. Nat Hum Behav. 2021 Apr;5(4):416–30.

4. Christensen GT, Mortensen EL, Christensen K, Osler M. Intelligence in young adulthood and cause-specific mortality in the Danish Conscription Database – A cohort study of 728,160 men. Intelligence. 2016 Nov 1;59:64–71.

5. Calvin CM, Batty GD, Der G, Brett CE, Taylor A, Pattie A, et al. Childhood intelligence in relation to major causes of death in 68 year follow-up: prospective population study. BMJ. 2017 Jun 28;357:j2708.

6. Deary IJ, Harris SE, Hill WD. What genome-wide association studies reveal about the association between intelligence and physical health, illness, and mortality. Curr Opin Psychol. 2019 Jun;27:6–12.

7. Luciano M, Batty GD, McGilchrist M, Linksted P, Fitzpatrick B, Jackson C, et al. Shared genetic aetiology between cognitive ability and cardiovascular disease risk factors: Generation Scotland’s Scottish family health study. Intelligence. 2010 May;38(3):304–13.

8. Mackenbach JP. The persistence of health inequalities in modern welfare states: The explanation of a paradox. Social Science & Medicine. 2012 Aug 1;75(4):761–9.

9. Arnett DK, Blumenthal RS, Albert MA, Buroker AB, Goldberger ZD, Hahn EJ, et al. 2019 ACC/AHA Guideline on the Primary Prevention of Cardiovascular Disease: A Report of the American College of Cardiology/American Heart Association Task Force on Clinical Practice Guidelines. Circulation. 2019 Sep 10;140(11):e596–646.

10. Richards M, Black S, Mishra G, Gale CR, Deary IJ, Batty DG. IQ in childhood and the metabolic syndrome in middle age: Extended follow-up of the 1946 British Birth Cohort Study. Intelligence. 2009 Nov 1;37(6):567–72.

11. Christensen GT, Rozing MP, Mortensen EL, Christensen K, Osler M. Young adult cognitive ability and subsequent major depression in a cohort of 666,804 Danish men. J Affect Disord. 2018 Aug 1;235:162–7.

12. Gale CR, Booth T, Starr JM, Deary IJ. Intelligence and socioeconomic position in childhood in relation to frailty and cumulative allostatic load in later life: the Lothian Birth Cohort 1936. J Epidemiol Community Health. 2016 Jun;70(6):576–82.

13. Yusuf S, Hawken S, Ounpuu S, Dans T, Avezum A, Lanas F, et al. Effect of potentially modifiable risk factors associated with myocardial infarction in 52 countries (the INTERHEART study): case-control study. Lancet. 2004 Sep 11;364(9438):937–52.

14. Lynch J, Davey Smith G, Harper S, Bainbridge K. Explaining the social gradient in coronary heart disease: comparing relative and absolute risk approaches. J Epidemiol Community Health. 2006 May;60(5):436–41.

15. Ariansen I, Graff-Iversen S, Stigum H, Strand BH, Wills AK, Næss Ø. Do repeated risk factor measurements influence the impact of education on cardiovascular mortality? Heart. 2015 Dec;101(23):1889–94.

16. Kuh D, Ben-Shlomo Y. Pre-adult influences on cardiovascular disease. In: Kuh D, Ben Shlomo Y, Ezra S, editors. A Life Course Approach to Chronic Disease Epidemiology [Internet]. Oxford University Press; 2004 [cited 2023 Dec 20]. p. 0. Available from: 10.1093/acprof:oso/9780198578154.003.0003

17. Davey Smith G, Hyppönen E, Power C, Lawlor DA. Offspring birth weight and parental mortality: prospective observational study and meta-analysis. American Journal of Epidemiology. 2007 Jul 15;166(2):160–9.

18. Kjøllesdal MKR, Carslake D, Davery Smith G, Shaikh F, Næss. The role of family factors in the association between early adulthood BMI and risk of cardiovascular disease. An intergenerational study of BMI in early adulthood and cardiovascular mortality in parents, aunts and uncles. International Journal of Obesity 2021 46:1. 2021 Oct 14;46(1):228–34.

19. Davey Smith G. Assessing Intrauterine Influences on Offspring Health Outcomes: Can Epidemiological Studies Yield Robust Findings? Basic & Clinical Pharmacology & Toxicology. 2008;102(2):245–56.

20. Fadum EA, Strand LAA, Rudvin I, Hæreid ML, Borud EK. The Norwegian Armed Forces Health Registry conscription board health examinations 1968–2018. Scandinavian Journal of Public Health [Internet]. 2020 [cited 2021 Nov 24]; Available from: https://pubmed.ncbi.nlm.nih.gov/32466714/

21. Bjartveit K, Foss OP, Gjervig T. The cardiovascular disease study in Norwegian counties. Results from first screening. Acta Medica Scandinavica, Supplement. 1983;675:1–184.

22. Tverdal A, Hjellvik V, Selmer R. Heart rate and mortality from cardiovascular causes: a 12 year follow-up study of 379 843 men and women aged 40-45 years. European Heart Journal. 2008 May 30;29(22):2772–81.

23. Naess O, Sogaard AJ, Arnesen E, Beckstrom AC, Bjertness E, Engeland A, et al. Cohort Profile: Cohort of Norway (CONOR). International Journal of Epidemiology. 2008 Jun 1;37(3):481–5.

24. Benson N, Hulac DM, Kranzler JH. Independent examination of the Wechsler Adult Intelligence Scale-Fourth Edition (WAIS-IV): what does the WAIS-IV measure? Psychological assessment. 2010 Mar;22(1):121–30.

25. Burke HR. Raven’s progressive matrices: a review and critical evaluation. The Journal of genetic psychology. 1958;93(2):199–228.

26. Hart CL, Taylor MD, Davey Smith G, Whalley LJ, Starr JM, Hole DJ, et al. Childhood IQ and cardiovascular disease in adulthood: prospective observational study linking the Scottish Mental Survey 1932 and the Midspan studies. Social Science & Medicine. 2004 Nov 1;59(10):2131–8.

27. Modig-Wennerstad K, Silventoinen K, Batty D, Tynelius P, Bergman L, Rasmussen F. Association between offspring intelligence and parental mortality: a population-based cohort study of one million Swedish men and their parents. Journal of Epidemiology & Community Health. 2008 Aug 1;62(8):722–7.

28. Batty GD, Mortensen EL, Nybo Andersen AM, Osler M. Childhood intelligence in relation to adult coronary heart disease and stroke risk: evidence from a Danish birth cohort study. Paediatric and Perinatal Epidemiology. 2005;19(6):452–9.

29. Sabia S, Guéguen A, Marmot MG, Shipley MJ, Ankri J, Singh-Manoux A. Does cognition predict mortality in midlife? Results from the Whitehall II cohort study. Neurobiology of aging. 2010 Apr;31(4):688.

30. Fraser A, Almqvist C, Larsson H, Långström N, Lawlor DA. Maternal diabetes in pregnancy and offspring cognitive ability: sibling study with 723,775 men from 579,857 families. Diabetologia. 2014;57(1):102–9.

31. Corrêa ML, Soares PSM, da Silva BGC, Wehrmeister F, Horta BL, Menezes AMB. Maternal smoking during pregnancy and intelligence quotient in offspring: A systematic review and meta-analysis. NeuroToxicology. 2021 Jul 1;85:99–114.

32. Kong A, Thorleifsson G, Frigge ML, Vilhjalmsson BJ, Young AI, Thorgeirsson TE, et al. The nature of nurture: Effects of parental genotypes. Science. 2018 Jan 26;359(6374):424–8.

33. Howe LJ, Nivard MG, Morris TT, Hansen AF, Rasheed H, Cho Y, et al. Within-sibship genome-wide association analyses decrease bias in estimates of direct genetic effects. Nat Genet. 2022 May;54(5):581–92.

34. Plomin R, Deary IJ. Genetics and intelligence differences: five special findings. Molecular Psychiatry. 2015 Feb 5;20(1):98–108.

35. Sunde HF, Eftedal NH, Cheesman R, Corfield EC, Kleppesto TH, Seierstad AC, et al. Genetic similarity between relatives provides evidence on the presence and history of assortative mating. Nat Commun. 2024 Mar 26;15(1):2641.

36. Border R, Athanasiadis G, Buil A, Schork AJ, Cai N, Young AI, et al. Cross-trait assortative mating is widespread and inflates genetic correlation estimates. Science. 2022 Nov 11;378(6621):754–61.

37. Watts GF, Gidding SS, Hegele RA, Raal FJ, Sturm AC, Jones LK, et al. International Atherosclerosis Society guidance for implementing best practice in the care of familial hypercholesterolaemia. Nat Rev Cardiol. 2023 Dec;20(12):845–69.

