## Supplementary tables for "Cognitive ability in offspring conscripts and cardiovascular disease risk in extended family members: assessing the impact of modifiable risk factors on familial risk"

**Supplementary table 1. Hazard ratios for cardiovascular and all-cause mortality among the parental generation per standard deviation (SD) unit of offspring cognitive ability (full-sample).** Hazard ratios (HR) and their corresponding 95% confidence intervals are shown for mothers, fathers, aunts/uncles (A/U) and A/U partners.

|  |  | **n** | **Model 1**  **Birth year + sex** | **Model 2**  **M1 + education** |
| --- | --- | --- | --- | --- |
| CVD mortality | Mothers | 614 650 | 0.75 (0.73, 0.77) | 0.84 (0.82, 0.86) |
|  | Fathers | 561 889 | 0.82 (0.80, 0.83) | 0.89 (0.87, 0.90) |
|  | A/U | 1 016 127 | 0.87 (0.86, 0.89) | 0.94 (0.92, 0.95) |
|  | A/U partners | 869 711 | 0.91 (0.90, 0.93) | 0.96 (0.94, 0.97) |
| All-cause mortality | Mothers | 614 650 | 0.85 (0.83, 0.84) | 0.90 (0.89, 0.91) |
|  | Fathers | 561 889 | 0.84 (0.83, 0.84) | 0.91 (0.90, 0.91) |
|  | A/U | 1 016 127 | 0.91 (0.90, 0.92) | 0.96 (0.95, 0.97) |
|  | A/U partners | 869 711 | 0.93 (0.92, 0.93) | 0.96 (0.96, 0.97) |

Note: Each SD unit is equal to 1.75 stanine or 13.13 intelligence quotient units. Education is measured in years of attained education. Abbreviations and symbols: n, number of observations; M1, model 1

**Supplementary table 2. Hazard ratio for CVD mortality among the parental generation in relation to offspring cognitive ability (CA) in the sub-sample of relatives with measured CVD risk factors.** Hazard ratios (HR) and their corresponding 95% confidence intervals are shown for mothers, fathers, aunts/uncles (A/U), and A/U partners.

|  | **n** | **2** | **3** | **4** | **5** | **6** | **7** | **8** | **9** | **Per stanine unit** |
| --- | --- | --- | --- | --- | --- | --- | --- | --- | --- | --- |
| **Mothers** |  |  |  |  |  |  |  |  |  |  |
| Model 1: Age + sex | 235 475 | 0.81 (0.61, 1.07) | 0.72 (0.55, 0.92) | 0.61 (0.48, 0.79) | 0.51 (0.40, 0.66) | 0.44 (0.34, 0.57) | 0.39 (0.30, 0.52) | 0.35 (0.26, 0.47) | 0.32 (0.22, 0.47) | 0.86 (0.84, 0.89) |
| Model 2: M1 + educ. | 235 475 | 0.83 (0.63, 1.10) | 0.77 (0.60, 1.00) | 0.70 (0.54, 0.91) | 0.62 (0.49, 0.81) | 0.56 (0.44, 0.73) | 0.54 (0.41, 0.71) | 0.51 (0.37, 0.69) | 0.50 (0.34, 0.74) | 0.91 (0.89, 0.94) |
| Model 3: M1 + CVD | 235 475 | 0.85 (0.64, 1.13) | 0.79 (0.61, 1.02) | 0.74 (0.57, 0.95) | 0.67 (0.52, 0.86) | 0.61 (0.47, 0.79) | 0.59 (0.45, 0.78) | 0.57 (0.42, 0.77) | 0.57 (0.38, 0.84) | 0.93 (0.90, 0.95) |
| Model 4: Full model | 235 475 | 0.87 (0.65, 1.15) | 0.82 (0.63, 1.06) | 0.78 (0.60, 1.00) | 0.72 (0.56, 0.93) | 0.68 (0.52, 0.88) | 0.67 (0.51, 0.88) | 0.66 (0.48, 0.89) | 0.67 (0.45, 1.00) | 0.95 (0.92, 0.98) |
| **Fathers** |  |  |  |  |  |  |  |  |  |  |
| Model 1: Age + sex | 235 475 | 0.97 (0.79, 1.19) | 0.89 (0.74, 1.08) | 0.80 (0.66, 0.96) | 0.69 (0.57, 0.83) | 0.61 (0.51, 0.74) | 0.54 (0.45, 0.66) | 0.54 (0.44, 0.67) | 0.51 (0.40, 0.66) | 0.90 (0.88, 0.92) |
| Model 2: M1 + educ. | 235 475 | 0.98 (0.80, 1.20) | 0.93 (0.77, 1.12) | 0.86 (0.71, 1.04) | 0.77 (0.64, 0.93) | 0.72 (0.60, 0.87) | 0.66 (0.54, 0.81) | 0.69 (0.56, 0.85) | 0.68 (0.53, 0.88) | 0.94 (0.92, 0.95) |
| Model 3: M1 + CVD | 235 475 | 0.99 (0.81, 1.22) | 0.94 (0.78, 1.14) | 0.88 (0.73, 1.06) | 0.81 (0.67, 0.97) | 0.77 (0.63, 0.93) | 0.71 (0.58, 0.87) | 0.74 (0.60, 0.92) | 0.75 (0.58, 0.96) | 0.95 (0.93, 0.96) |
| Model 4: Full model | 235 475 | 0.99 (0.81, 1.22) | 0.96 (0.79, 1.16) | 0.90 (0.75, 1.09) | 0.84 (0.70, 1.02) | 0.82 (0.67, 0.99) | 0.77 (0.63, 0.93) | 0.82 (0.66, 1.01) | 0.84 (0.65, 1.08) | 0.96 (0.94, 0.98) |
| **A/U** |  |  |  |  |  |  |  |  |  |  |
| Model 1: Age + sex | 235 475 | 0.91 (0.71, 1.18) | 0.91 (0.72, 1.16) | 0.91 (0.72, 1.15) | 0.85 (0.67, 1.08) | 0.72 (0.57, 0.92) | 0.74 (0.58, 0.94) | 0.64 (0.50, 0.83) | 0.77 (0.57, 1.03) | 0.95 (0.93, 0.97) |
| Model 2: M1 + educ. | 235 475 | 0.94 (0.73, 1.20) | 0.97 (0.76, 1.23) | 1.00 (0.79, 1.27) | 0.97 (0.77, 1.23) | 0.86 (0.68, 1.09) | 0.91 (0.71, 1.16) | 0.82 (0.63, 1.07) | 1.04 (0.78, 1.40) | 0.98 (0.96, 1.00) |
| Model 3: M1 + CVD | 235 475 | 0.92 (0.72, 1.19) | 0.95 (0.75, 1.21) | 0.99 (0.78, 1.25) | 0.97 (0.76, 1.22) | 0.85 (0.67, 1.08) | 0.88 (0.69, 1.13) | 0.80 (0.62, 1.04) | 1.00 (0.74, 1.34) | 0.98 (0.96, 1.00) |
| Model 4: Full model | 235 475 | 0.94 (0.73, 1.21) | 0.99 (0.78, 1.25) | 1.04 (0.82, 1.31) | 1.03 (0.82, 1.30) | 0.92 (0.73,1.17) | 0.98 (0.77, 1.26) | 0.90 (0.69, 1.18) | 1.16 (0.87, 1.56) | 1.00 (0.97, 1.02) |
| **A/U partners** |  |  |  |  |  |  |  |  |  |  |
| Model 1: Age + sex | 235 475 | 1.13 (0.91, 1.39) | 1.06 (0.86, 1.30) | 1.00 (0.82, 1.23) | 0.94 (0.71, 1.15) | 0.90 (0.73, 1.11) | 0.83 (0.67, 1.03) | 0.81 (0.65, 1.02) | 0.74 (0.57, 0.96) | 0.95 (0.93, 0.97) |
| Model 2: M1 + educ. | 235 475 | 1.14 (0.92, 1.40) | 1.09 (0.89, 1.35) | 1.06 (0.87, 1.31) | 1.02 (0.83, 1.25) | 1.01 (0.82, 1.24) | 0.96 (0.78, 1.20) | 0.96 (0.76, 1.20) | 0.90 (0.70, 1.17) | 0.97 (0.96, 0.99) |
| Model 3: M1 + CVD | 235 475 | 1.15 (0.93, 0.42) | 1.10 (0.89, 1.35) | 1.05 (0.86, 1.29) | 1.02 (0.83, 1.25) | 1.02 (0.82, 1.25) | 0.96 (0.78, 1.19) | 0.97 (0.77, 1.21) | 0.92 (0.71, 1.19) | 0.98 (0.96, 0.99) |
| Model 4: Full model | 235 475 | 1.15 (0.93, 0.42) | 1.12 (0.91, 1.37) | 1.09 (0.88, 1.33) | 1.06 (0.87, 1.30) | 1.06 (0.87, 1.31) | 1.02 (0.83, 1.27) | 1.05 (0.83, 1.31) | 1.01 (0.78, 1.31) | 0.99 (0.97, 1.00) |

**Note:** Exposure was analysed as categorical (see Methods section). Reference category was stanine score 1. Each stanine score is equal to 7.5 intelligence quotient points. Abbreviations and symbols: n, number of observations; M1, model 1; educ., education (years of attained education); CVD, cardiovascular disease risk factors (body mass index, non-fasting cholesterol, systolic blood pressure, smoking).

**Supplementary table 3. Hazard ratio for all-cause mortality among the parental generation in relation to offspring cognitive ability (CA) in the sub-sample of relatives with measured CVD risk factors.** Hazard ratios (HR) and their corresponding 95% confidence intervals are shown for mothers, fathers, aunts/uncles (A/U), and A/U partners.

|  | **n** | **2** | **3** | **4** | **5** | **6** | **7** | **8** | **9** | **Per stanine unit** |
| --- | --- | --- | --- | --- | --- | --- | --- | --- | --- | --- |
| **Mothers** |  |  |  |  |  |  |  |  |  |  |
| Model 1: Age + sex | 235 475 | 0.90 (0.81, 1.00) | 0.81 (0.73, 0.90) | 0.71 (0.65, 0.79) | 0.66 (0.60, 0.73) | 0.60 (0.55, 0.67) | 0.55 (0.50, 0.61) | 0.52 (0.46, 0.58) | 0.50 (0.43, 0.57) | 0.91 (0.90, 0.92) |
| Model 2: M1 + educ. | 235 475 | 0.92 (0.82, 1.02) | 0.86 (0.77, 0.95) | 0.78 (0.70, 0.86) | 0.75 (0.68, 0.83) | 0.72 (0.65, 0.79) | 0.68 (0.61, 0.76) | 0.66 (0.59, 0.74) | 0.67 (0.58, 0.76) | 0.95 (0.93, 0.96) |
| Model 3: M1 + CVD | 235 475 | 0.92 (0.83, 1.03) | 0.86 (0.77, 0.95) | 0.79 (0.71, 0.87) | 0.77 (0.69, 0.85) | 0.74 (0.67, 0.82) | 0.70 (0.63, 0.78) | 0.69 (0.61, 0.77) | 0.70 (0.61, 0.80) | 0.95 (0.95, 0.96) |
| Model 4: Full model | 235 475 | 0.93 (0.84, 1.04) | 0.88 (0.79, 0.97) | 0.82 (0.74, 0.91) | 0.81 (0.73, 0.90) | 0.79 (0.72, 0.88) | 0.77 (0.69, 0.86) | 0.76 (0.68, 0.86) | 0.79 (0.70, 0.90) | 0.97 (0.96, 0.98) |
| **Fathers** |  |  |  |  |  |  |  |  |  |  |
| Model 1: Age + sex | 235 475 | 0.93 (0.84, 1.03) | 0.88 (0.80, 0.97) | 0.79 (0.72, 0.87) | 0.72 (0.65, 0.79) | 0.64 (0.58, 0.70) | 0.62 (0.56, 0.68) | 0.57 (0.51, 0.64) | 0.54 (0.47, 0.61) | 0.92 (0.91, 0.93) |
| Model 2: M1 + educ. | 235 475 | 0.94 (0.84, 1.04) | 0.91 (0.83, 1.01) | 0.85 (0.77, 0.94) | 0.80 (0.72, 0.88) | 0.74 (0.67, 0.81) | 0.74 (0.67, 0.82) | 0.71 (0.64, 0.79) | 0.70 (0.61, 0.79) | 0.95 (0.94, 0.96) |
| Model 3: M1 + CVD | 235 475 | 0.95 (0.85, 1.05) | 0.92 (0.83, 1.01) | 0.86 (0.78, 0.94) | 0.81 (0.73, 0.89) | 0.75 (0.68, 0.83) | 0.75 (0.68, 0.83) | 0.72 (0.65, 0.80) | 0.70 (0.62, 0.80) | 0.95 (0.94, 0.96) |
| Model 4: Full model | 235 475 | 0.95 (0.86, 1.06) | 0.94 (0.85, 1.03) | 0.88 (0.80, 0.97) | 0.85 (0.77, 0.94) | 0.80 (0.73, 0.89) | 0.82 (0.74, 0.91) | 0.80 (0.72, 0.90) | 0.80 (0.71, 0.91) | 0.97 (0.96, 0.98) |
| **A/U** |  |  |  |  |  |  |  |  |  |  |
| Model 1: Age + sex | 235 475 | 0.95 (0.85, 1.07) | 0.92 (0.82, 1.02) | 0.91 (0.82, 1.01) | 0.86 (0.77, 0.95) | 0.78 (0.70, 0.87) | 0.77 (0.69, 0.86) | 0.73 (0.65, 0.82) | 0.70 (0.61, 0.80) | 0.95 (0.95, 0.96) |
| Model 2: M1 + educ. | 235 475 | 0.97 (0.86, 1.09) | 0.96 (0.86, 1.07) | 0.97 (0.87, 1.08) | 0.94 (0.85, 1.05) | 0.89 (0.80, 0.99) | 0.90 (0.80, 1.01) | 0.87 (0.77, 0.98) | 0.87 (0.75, 1.00) | 0.98 (0.97, 0.99) |
| Model 3: M1 + CVD | 235 475 | 0.95 (0.85, 1.07) | 0.94 (0.86, 1.05) | 0.95 (0.86, 1.06) | 0.93 (0.84, 1.03) | 0.87 (0.78, 0.97) | 0.88 (0.79, 0.98) | 0.85 (0.76, 0.96) | 0.83 (0.72, 0.95) | 0.98 (0.97, 0.99) |
| Model 4: Full model | 235 475 | 0.96 (0.86, 1.08) | 0.96 (0.86, 1.07) | 0.99 (0.89, 1.10) | 0.97 (0.88, 1.08) | 0.93 (0.83, 1.03) | 0.95 (0.85, 1.06) | 0.93 (0.83, 1.04) | 0.93 (0.81, 1.06) | 0.99 (0.98, 1.00) |
| **A/U partners** |  |  |  |  |  |  |  |  |  |  |
| Model 1: Age + sex | 235 475 | 0.94 (0.86, 1.02) | 0.95 (0.87, 1.04) | 0.90 (0.83, 0.98) | 0.86 (0.79, 0.93) | 0.83 (0.76, 0.90) | 0.79 (0.73, 0.87) | 0.77 (0.71, 0.85) | 0.69 (0.62, 0.77) | 0.96 (0.95, 0.97) |
| Model 2: M1 + educ. | 235 475 | 0.95 (0.86, 1.03) | 0.97 (0.89, 1.06) | 0.94 (0.86, 1.02) | 0.91 (0.84, 0.99) | 0.90 (0.82, 0.97) | 0.87 (0.80, 0.95) | 0.87 (0.79, 0.95) | 0.80 (0.72, 0.89) | 0.98 (0.97, 0.99) |
| Model 3: M1 + CVD | 235 475 | 0.95 (0.87, 1.04) | 0.98 (0.90, 1.06) | 0.94 (0.87, 1.02) | 0.92 (0.84, 1.00) | 0.91 (0.83, 0.99) | 0.88 (0.80, 0.97) | 0.88 (0.80, 0.97) | 0.82 (0.73, 0.91) | 0.98 (0.97, 0.99) |
| Model 4: Full model | 235 475 | 0.95 (0.87, 1.04) | 0.99 (0.91, 1.07) | 0.96 (0.88, 1.04) | 0.94 (0.86, 1.02) | 0.94 (0.86, 1.02) | 0.92 (0.84, 1.01) | 0.93 (0.84, 1.01) | 0.87 (0.78, 0.97) | 0.99 (0.98, 1.00) |

**Note:** Exposure was analysed as categorical (see Methods section). Reference category was stanine score 1. Each stanine score is equal to 7.5 intelligence quotient points. Abbreviations and symbols: n, number of observations; M1, model 1; educ., education (years of attained education); CVD, cardiovascular disease risk factors (body mass index, non-fasting cholesterol, systolic blood pressure, smoking).

**Supplementary table 4. Hazard ratio for CVD and all-cause and CVD mortality among the parental generation in relation to offspring cognitive ability (CA) in the full sample.** Hazard ratios (HR) and their corresponding 95% confidence intervals are shown for mothers, fathers, aunts/uncles (A/U), and A/U partners.

|  |  | **n** | **2** | **3** | **4** | **5** | **6** | **7** | **8** | **9** | **Per stanine unit** |
| --- | --- | --- | --- | --- | --- | --- | --- | --- | --- | --- | --- |
| CVD mortality | **Mothers** |  |  |  |  |  |  |  |  |  |  |
|  | Model 1: Age + sex | 235 475 | 0.77 (0.67, 0.89) | 0.65 (0.57, 0.75) | 0.59 (0.51, 0.67) | 0.48 (0.42, 0.55) | 0.40 (0.35, 0.46) | 0.35 (0.30, 0.41) | 0.30 (0.25, 0.35) | 0.26 (0.21, 0.32) | 0.85 (0.84, 0.86) |
|  | Model 2: M1 + educ. | 235 475 | 0.80 (0.69, 0.93) | 0.71 (0.62, 0.81) | 0.68 (0.59, 0.77) | 0.59 (0.52, 0.68) | 0.53 (0.46, 0.61) | 0.50 (0.43, 0.58) | 0.45 (0.38, 0.54) | 0.43 (0.35, 0.53) | 0.90 (0.89, 0.92) |
|  | **Fathers** |  |  |  |  |  |  |  |  |  |  |
|  | Model 1: Age + sex | 235 475 | 0.84 (0.76, 0.93) | 0.71 (0.65, 0.78) | 0.66 (0.60, 0.72) | 0.57 (0.52, 0.62) | 0.51 (0.46, 0.56) | 0.45 (0.41, 0.50) | 0.42 (0.38, 0.47) | 0.40 (0.35, 0.46) | 0.89 (0.88, 0.90) |
|  | Model 2: M1 + educ. | 235 475 | 0.86 (0.78, 0.95) | 0.75 (0.68, 0.82) | 0.73 (0.67, 0.80) | 0.67 (0.61, 0.73) | 0.63 (0.57, 0.69) | 0.58 (0.53, 0.64) | 0.57 (0.52, 0.64) | 0.57 (0.51, 0.65) | 0.93 (0.92, 0.94) |
|  | **A/U** |  |  |  |  |  |  |  |  |  |  |
|  | Model 1: Age + sex | 235 475 | 0.88 (0.78, 1.00) | 0.86 (0.77, 0.97) | 0.80 (0.71, 0.89) | 0.74 (0.66, 0.83) | 0.66 (0.59, 0.74) | 0.63 (0.56, 0.72) | 0.59 (0.52, 0.67) | 0.53 (0.46, 0.62) | 0.93 (0.92, 0.94) |
|  | Model 2: M1 + educ. | 235 475 | 0.91 (0.80, 1.03) | 0.92 (0.81, 1.03) | 0.88 (0.79, 0.99) | 0.86 (0.76, 0.96) | 0.79 (0.70, 0.88) | 0.79 (0.70, 0.89) | 0.76 (0.67, 0.87) | 0.73 (0.62, 0.85) | 0.96 (0.95, 0.97) |
|  | **A/U partners** |  |  |  |  |  |  |  |  |  |  |
|  | Model 1: Age + sex | 235 475 | 0.96 (0.86, 1.07) | 0.91 (0.82, 1.01) | 0.82 (0.74, 0.91) | 0.78 (0.71, 0.87) | 0.76 (0.69, 0.84) | 0.73 (0.66, 0.82) | 0.68 (0.60, 0.76) | 0.64 (0.56, 0.73) | 0.95 (0.94, 0.96) |
|  | Model 2: M1 + educ. | 235 475 | 0.97 (0.87, 1.08) | 0.94 (0.85, 1.05) | 0.87 (0.79, 0.97) | 0.86 (0.77, 0.95) | 0.86 (0.77, 0.95) | 0.85 (0.76, 0.95) | 0.81 (0.72, 0.91) | 0.79 (0.70, 0.91) | 0.97 (0.97, 0.98) |
| All-cause  mortality | **Mothers** |  |  |  |  |  |  |  |  |  |  |
|  | Model 1: Age + sex | 235 475 | 0.88 (0.82, 0.93) | 0.78 (0.73, 0.83) | 0.70 (0.66, 0.75) | 0.63 (0.59, 0.67) | 0.57 (0.54, 0.60) | 0.52 (0.49, 0.55) | 0.47 (0.44, 0.50) | 0.46 (0.42, 0.49) | 0.90 (0.90, 0.91) |
|  | Model 2: M1 + educ. | 235 475 | 0.90 (0.85, 0.96) | 0.83 (0.80, 0.88) | 0.78 (0.74, 0.82) | 0.73 (0.69, 0.77) | 0.69 (0.65, 0.73) | 0.66 (0.62, 0.70) | 0.63 (0.58, 0.67) | 0.64 (0.59, 0.69) | 0.94 (0.94, 0.95) |
|  | **Fathers** |  |  |  |  |  |  |  |  |  |  |
|  | Model 1: Age + sex | 235 475 | 0.91 (0.86, 0.96) | 0.81 (0.77, 0.85) | 0.73 (0.69, 0.76) | 0.64 (0.61, 0.67) | 0.58 (0.55, 0.61) | 0.54 (0.51, 0.60) | 0.50 (0.47, 0.53) | 0.47 (0.44, 0.50) | 0.91 (0.90, 0.91) |
|  | Model 2: M1 + educ. | 235 475 | 0.93 (0.87, 0.98) | 0.85 (0.81, 0.90) | 0.80 (0.76, 0.84) | 0.74 (0.70, 0.78) | 0.70 (0.67, 0.74) | 0.68 (0.65, 0.72) | 0.66 (0.63, 0.70) | 0.65 (0.61, 0.70) | 0.95 (0.94, 0.95) |
|  | **A/U** |  |  |  |  |  |  |  |  |  |  |
|  | Model 1: Age + sex | 235 475 | 0.96 (0.90, 1.02) | 0.92 (0.87, 0.97) | 0.86 (0.81, 0.91) | 0.82 (0.78, 0.87) | 0.77 (0.73, 0.81) | 0.73 (0.69, 0.78) | 0.70 (0.66, 0.75) | 0.67 (0.62, 0.72) | 0.95 (0.94, 0.95) |
|  | Model 2: M1 + educ. | 235 475 | 0.98 (0.92, 1.04) | 0.96 (0.91, 1.02) | 0.93 (0.88, 0.99) | 0.92 (0.87, 0.97) | 0.89 (0.84, 0.94) | 0.87 (0.82, 0.92) | 0.86 (0.80, 0.91) | 0.85 (0.79, 0.91) | 0.98 (0.97, 0.98) |
|  | **A/U partners** |  |  |  |  |  |  |  |  |  |  |
|  | Model 1: Age + sex | 235 475 | 0.98 (0.93, 1.03) | 0.95 (0.90, 0.99) | 0.89 (0.85, 0.93) | 0.85 (0.81, 0.89) | 0.81 (0.77, 0.85) | 0.79 (0.76, 0.83) | 0.76 (0.72, 0.80) | 0.72 (0.68, 0.76) | 0.96 (0.95, 0.96) |
|  | Model 2: M1 + educ. | 235 475 | 0.99 (0.94, 1.04) | 0.97 (0.93, 1.02) | 0.93 (0.89, 0.97) | 0.91 (0.87, 0.95) | 0.89 (0.85, 0.93) | 0.89 (0.85, 0.93) | 0.87 (0.83, 0.92) | 0.85 (0.80, 0.90) | 0.98 (0.98, 0.98) |

**Note:** Exposure was analysed as categorical ordinal variable (see Methodology). Reference category was stanine score 1. Each stanine score is equal to 7.5 intelligence quotient points. Abbreviations and symbols: n, number of observations; M1, model 1; educ., education (years of attained education).
